# COVID-19 higher induced mortality in Chinese regions with lower air quality

**DOI:** 10.1101/2020.04.04.20053595

**Authors:** Riccardo Pansini, Davide Fornacca

## Abstract

COVID-19 has spread in all continents in a span of just over three months, escalating into a pandemic that poses several humanitarian as well as scientific challenges. We here investigated the geographical expansion of the infection and correlate it with the annual indexes of air quality observed from the Sentinel-5 satellite orbiting around China, Italy and the U.S.A. Controlling for population size, we find more viral infections in those areas afflicted by Carbon Monoxide (CO) and Nitrogen Dioxide (NO_2_). Higher mortality was also correlated with poor air quality, namely with high PM2.5, CO and NO_2_ values. In Italy, the correspondence between poor air quality and SARS-CoV-2 appearance and induced mortality was the starkest. Similar to smoking, people living in polluted areas are more vulnerable to SARS-CoV-2 infections and induced mortality. This further suggests the detrimental impact climate change will have on the trajectory of future epidemics.

**Significance:** We found a significant correlation between levels of air quality and COVID-19 spread and mortality in China, Italy and the United States. Despite the infection being still ongoing at a global level, these correlations are relatively robust not being influenced by varying population densities. Living in an area with low air quality seems to be a risk factor for becoming infected and dying from this new form of coronavirus.

## 1. Introduction

From the first detected **outbreak** of a new member of the coronavirus (CoV) family^1^ in Wuhan, Hubei Province, China^2,3^, SARS-CoV-2^4^ has rapidly spread around the world^5^, with governments and institutions showing mixed results in its effective containment^6^. Certain regions have been much more adversely impacted in terms of the rate of infections and mortality rates than others, and the full reasons for this are not yet clear. This paper shows preliminary, yet compelling evidence of a correlation between air pollution and incidence of COVID-19 in China, Italy and the United States.

**Air pollution** is notoriously known to cause health problems, and, in particular, respiratory diseases, to individuals exposed for longer than several days per year^7-10^. Moreover, pollutants in the air are a significant underlying contributors to the emergence of respiratory viral infections^11^. In particular, PM10 and PM2.5 have been linked to respiratory disease hospitalisations for pneumonia and chronic pulmonary diseases^12-18^. There is some further experimental evidence that emissions from diesel and coal affect the lungs, causing pathological immune response and inflammations^19,20^.

Airborne microorganisms can directly infect other people’s mucosae or travel further into the air and onto surfaces causing delayed infections. The **particles** of several pollutants such as PM and NO_2_ can act as a vector for the spread and extended survival in the air of bioaerosols^21-26^ including viruses^27-31^. A first hypothesis in this direction has arisen for COVID-19 in Northern Italy^32^ – granted that the viral load in a flying aggregate can be enough to cause morbidity.

Several **risk factors** have been implicated with the fast spread of the virus, including super spread events^33,34^. Its further spread to different countries has been attributed to air travellers^35-37,6,38-42^. A number of personal risk factors have further been implicated with higher morbidity and mortality rates of Covid-19, including male gender and smoking status. In particular, smoking has been associated with a higher morbidity and mortality of COVID-19 in men than in women^43^.

However, the very **first appearance** of the virus cannot be directly correlated with one of these predictors, since, like the other SARS coronaviruses, SARS-CoV-2 is alleged to have transferred host from the originating bats to humans^44^. However, it still appeared in a Chinese area affected by some of the highest air pollution in the world, and it showed a relatively high virulence there.

The strong **containment** measures adopted firstly by the Chinese government have necessarily biased the natural virus spread^45^, not allowing the virus to distribute evenly to polluted and non-polluted areas of the country. As a result, these measures have been highly effective^46^. This applies also to those other countries where similar effective containment measures were taken at the earlier stages of the outbreaks^47^. There is nevertheless mounting evidence that the outbreaks went undetected for weeks, as in the case of northern Italy, imputed to be as early as 1 January 2020 and not as the first case registered on 20 February 2020^48^. This latter element may mitigate the role that the containment measures had at containing the diffusion of the virus, leading to air pollution playing a more relevant role in the incidence of the virus.

Together with smoking, several others are the **predictor variables** ascribed to the incidence of COVID-19.

A high **population density** is the first predictor variable for the virus spread, but it cannot be a predictor for a higher virulence and a higher mortality^49^. For instance, in Italy^50^, the metropolis of Milan was not as affected as the inhabited surrounding areas with lower density^51^.

Another evident predictor variable is **transportation**. The surrounding areas of transport hubs such as airports and large train stations should witness the appearance of the virus earlier than other geographical areas and they act as transmission hubs^52-56^.

The temperate-climate **latitudes** have been identified as the probable areas to be mostly affected by COVID-19^57^ due to a limited exposure to UV light in winter. The sole temperature^58,59^ or humidity^60^ appear to play less of a role. Indeed, other human coronaviruses (HCoV-229E, HCoV-HKU1, HCoV-NL63, and HCoV-OC43) appear between December and April, and are undetectable in summer months in temperate regions, leading to winter seasonality behaviour.

One factor that has so far been overlooked is the role of **air pollution** in the spread and mortality rates of COVID-19. Air pollution has been shown to be strongly associated with high incidence of other respiratory infections^12,13,7,14,15,11,10,16-18^ and higher mortality rates^8,9^.

Here we investigate whether there is a correlation between air pollution and air-borne SARS-CoV-2 causing respiratory diseases in China, Italy^61^ and the United States. Our **hypotheses** were:

Hyp 1: Is there a higher incidence of COVID-19 in highly polluted areas?

Hyp 2: Is there a higher COVID-19 mortality rate in highly polluted areas?

## 2. Methods

For the time being, we renounced at performing a comparative study at a worldwide scale, due to significant differences in the coverage and compiling methodology of the COVID-19 infections and deaths among countries. Instead, we selected three countries particularly affected by the virus, and evaluated the potential correlation between air quality metrics and infections at the most detailed level of data available. Results for each country were analysed separately. China (including Taiwan, Hong Kong and Macau) was chosen because of its large size and now advanced stage of its epidemic, well into the recovery phase. The second choice was Italy, at the time of writing the most heavily affected country of the world, just passed the peak of contagion. The area with the largest number of infections and deaths in Italy is the Po Valley, which is also the foremost place of polluted air in Europe^62^. The third country investigated was the conterminous U.S.A., which currently has the highest number of cases worldwide, yet is still at an early stage of the pandemic due to its later arrival as compared to Asia and Europe.

Table 1 The datasets used for the viral and pollution analyses.

**Table 1.**
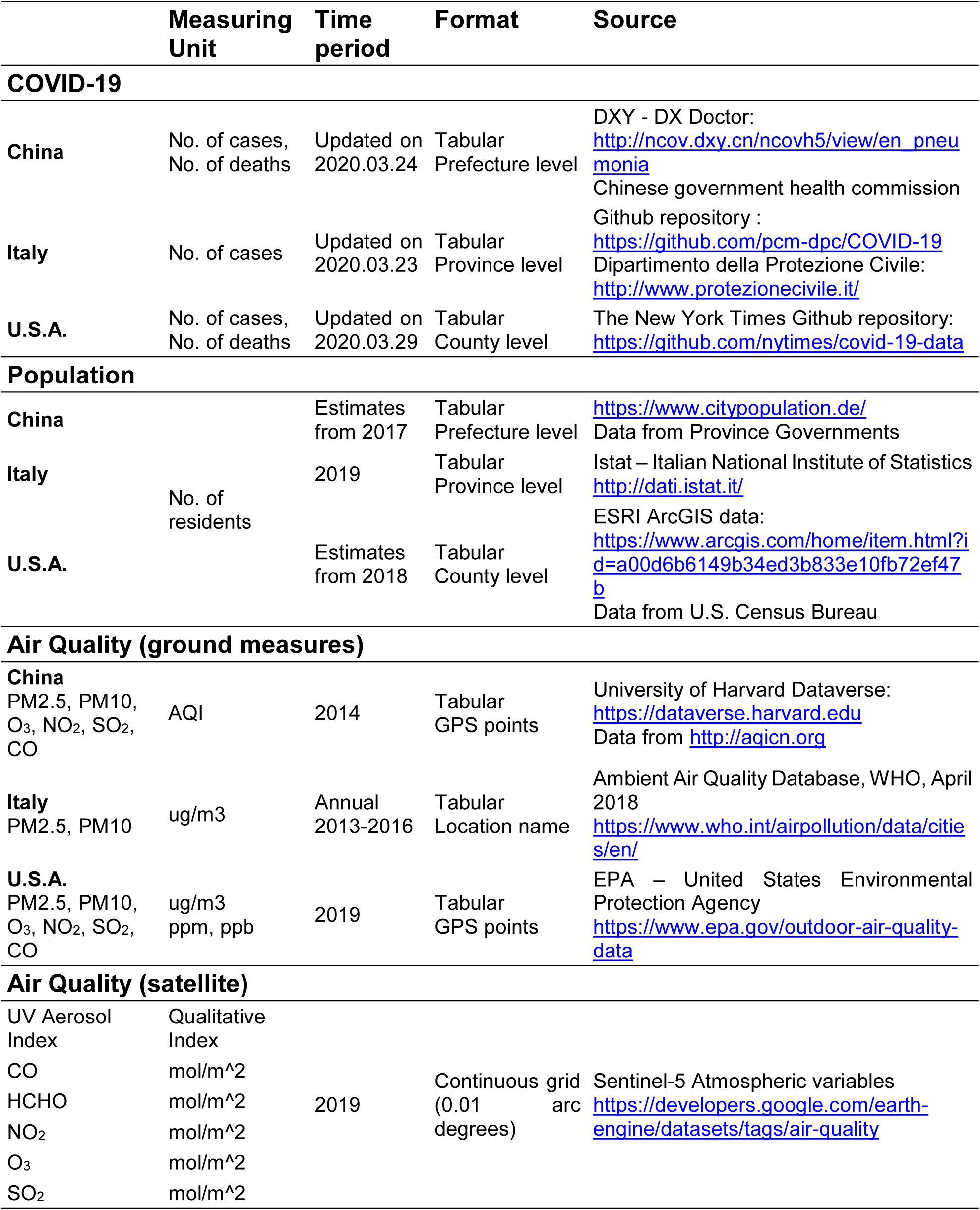
summarises the datasets used:

### 2.1 Data collection

The dataset was compiled at the second-level administrative subdivision, which corresponds to prefectures for China, provinces for Italy and counties for the U.S.A. Both cases and deaths due to COVID-19 were collected and normalised by population size (100,000 residents). Air quality information came from two kinds of sources: measured values from ground monitoring stations and Sentinel-5 satellite data. The latest available annual means of measured PM2.5 and PM10 values were retrieved from the World Health Organization Global Ambient Air Quality Database of 2018^63^ and used for the Italian case study because of extensive coverage. For China, we used aggregated monthly air quality data for the years 2014 to 2016, made available by the Center for Geographic Analysis Dataverse of the University of Harvard^64^. For the conterminous U.S.A., summary data on several pollutants for the year 2019 was retrieved from the United States Environmental Protection Agency^65^. To every administrative unit we assigned the air quality value from its related station. If more than one point fell within a given unit, the mean was calculated. These three datasets were used for the analysis but also to validate the air quality data derived from satellite imagery. The Sentinel-5 mission from the Copernicus program of the European Space Agency was launched in October 2017 and was specifically designed to provide coherent information on atmospheric variables for air quality, ozone and climate^66^. By means of the Google Earth Engine platform^67^ which provides open-access satellite data organized in time-series, we retrieved 6 different datasets derived from Sentinel-5 series. These variables were the UV Aerosol Index, Carbon Monoxide (CO), Formaldehyde (HCHO), Nitrogen Dioxide (NO_2_), Ozone (O_3_), and Sulphur Dioxide (SO_2_). Every time-series was processed to obtain the annual mean of the year 2019 and we calculated the mean of all grid cells covering every administrative unit.

Table 1 summarises the datasets used:

### 2.2 Data Analysis

Skewness and Kurtosis were calculated for each variable to evaluate the normality of the distributions. Pearson and Kendall correlation matrices were produced and the corresponding coefficients were analysed according to the distribution type. We were particularly interested in the following relationships:

- COVID-19 cases / 100,000 habitants vs. single air pollution variables;
- COVID-19 deaths / 100,000 habitants vs. single air pollution variables;
- Correlation between ground-measured pollution values vs. satellite-derived values. Satellite data should in fact be more advantageous than ground station data, because of their regular and continuous data acquisition, quasi-global coverage, and spatially consistent measurement methodology. On the other hand, ground stations offer real measures of single pollutants instead of deriving it from spectral information (satellites), however, they require more or less arbitrary estimations (such as interpolation) to fill spatial gaps.

Thematic maps for visual interpretation were produced to better highlight the potential air quality and COVID-19 distributions within the three assessed countries.

## 3. Results

The main dependent variables for our study, namely the number of COVID-19 cases per 100,000 inhabitants and the mortality rate (no. of deaths/no. of cases) presented both highly skewed distributions in the three countries that we analysed. However, by setting the reference value range between -1.96 and 1.96 for both skewness and kurtosis, several of the air quality variables showed fairly normal distributions (Table 2). For the following correlation results, we report both Pearson’s Rho and Kendall’s Tau coefficients, and highlight the most suitable one according to the distribution of the variables.

**Table 2.** Skewness and Kurtosis values for the distribution of the analysed variables.

|  | China |  | Italy |  | U.S.A. |  |
| --- | --- | --- | --- | --- | --- | --- |
|  | Skew. | Kurt. | Skew. | Kurt. | Skew. | Kurt. |
| **cases_100k** | 14.02 | 224.34 | 3.01 | 10.22 | 12.83 | 243.19 |
| **mortality** | 4.77 | 31.05 | - | - | 11.17 | 139.41 |
| **Aerosol_sat** | 0.63 | 1.51 | 1.72 | 5.28 | 0.68 | 0.63 |
| **CO_sat** | -0.91 | 0.18 | -1.07 | 2.63 | -1.84 | 3.85 |
| **HCHO_sat** | -0.63 | -0.27 | -1.00 | 2.84 | -0.33 | 0.23 |
| **NO<sub>2</sub>_sat** | 1.13 | 0.8 | 1.68 | 3.45 | 2.94 | 21.70 |
| **O<sub>3</sub>_sat** | 0.42 | -0.77 | -0.80 | -0.08 | -0.14 | -0.94 |
| **SO<sub>2</sub>_sat** | 1.29 | 2.77 | 4.89 | 30.16 | 0.73 | 4.06 |
| **PM25_ground** | 0 | 0.02 | 0.44 | -0.32 | -0.17 | 0.35 |
| **PM10_ground** | 0.94 | 1.73 | 0.70 | 0.14 | 1.25 | 2.69 |
| **CO_ground** | 1.15 | 1.85 | - | - | 0.29 | 1.01 |
| **NO<sub>2</sub>_ground** | 0.27 | -0.55 | - | - | 0.36 | -0.51 |
| **O<sub>3</sub>_ground** | 0.55 | 0.48 | - | - | -0.27 | 1.42 |
| **SO<sub>2</sub>_ground** | 1.08 | 0.67 | - | - | 8.87 | 103.49 |

**Graph 1.**
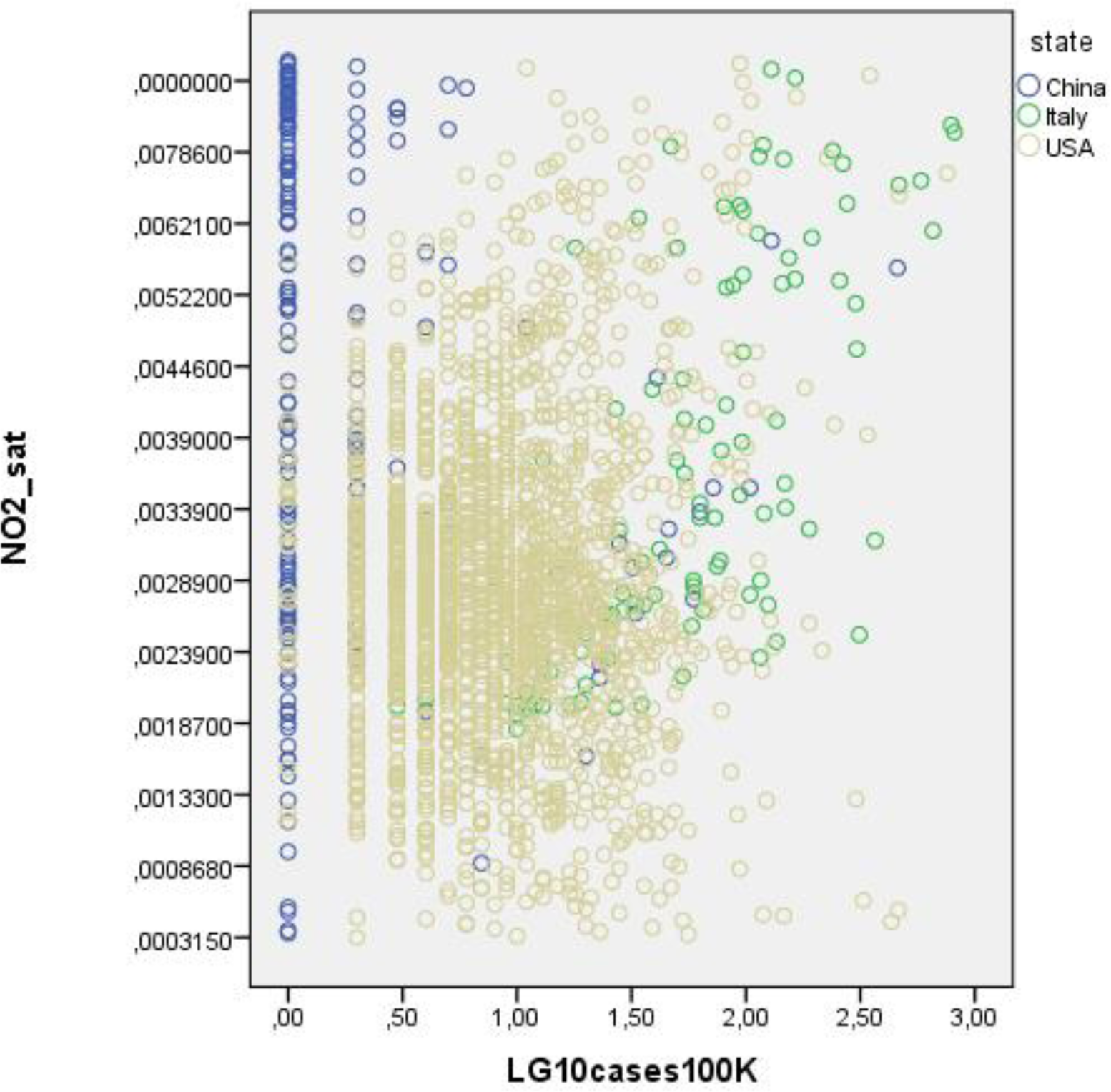
Scatter plot of COVID-19 total cases (controlled for population density) and satellite NO_2_ concentrations for the three countries. Compared to China and the U.S.A., Italy exhibits the most infections per capita correlated with increasing levels of air pollution.

### 3.1 Comparison of satellite-derived data with ground measures

**Figure 1.**
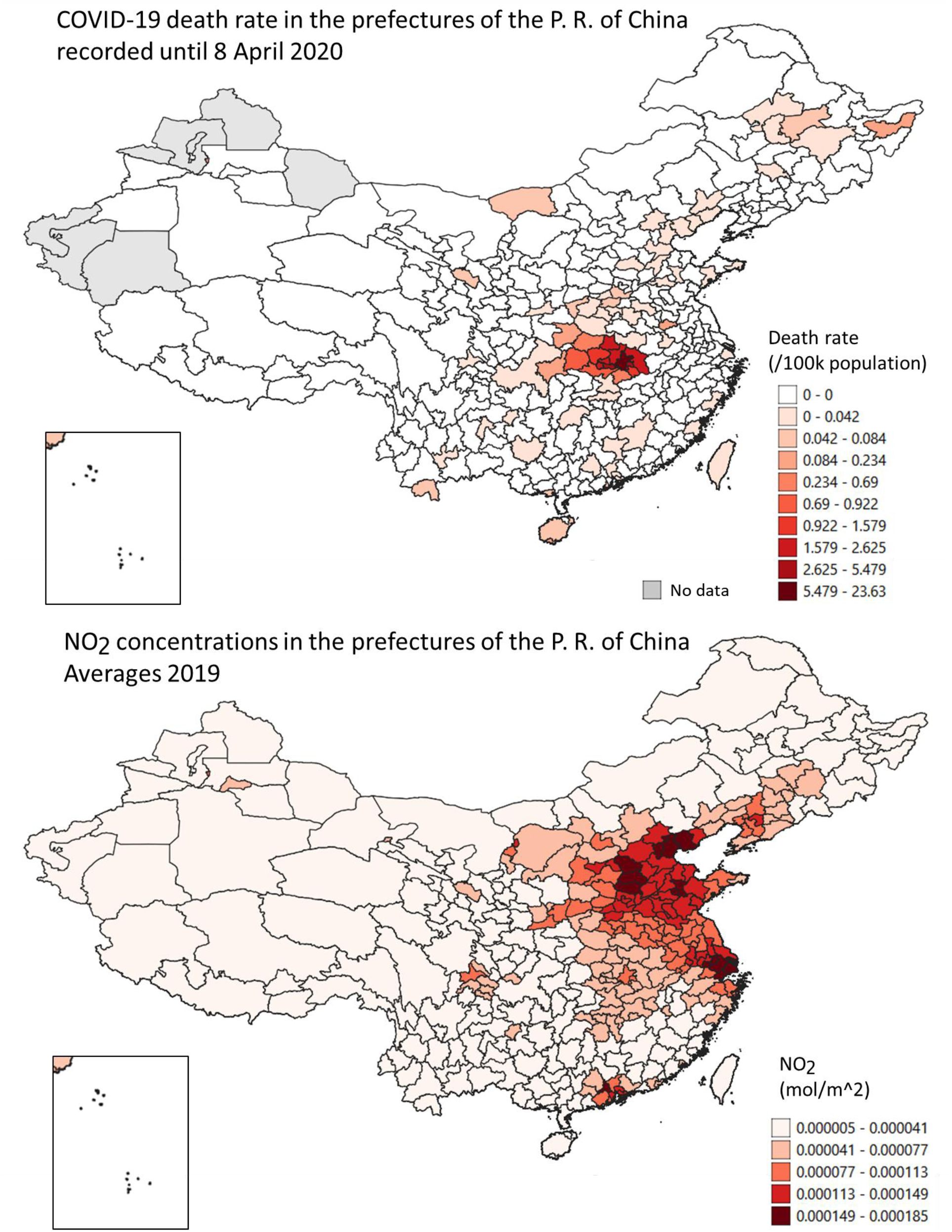
Map of COVID-19 total cases (controlled for population density) and NO_2_ concentrations in the P.R. of China.

The data collected for China and the U.S.A. allowed for a comparison between some of the satellite-derived air pollution variables with ground stations measures, namely CO, NO_2_, O_3_, and SO_2_ (Table 3). The strongest correlation was found in the NO_2_ values, both in China and in the U.S.A. Carbon Dioxide and SO_2_ also showed significant agreement in China, while in the U.S.A. no correlation was found for SO_2_. In both countries, the Ozone values did not show a significant correlation.

**Table 3.** Correlations between satellite-derived values and ground measures. Greyed cells indicate the more adequate statistical test according to data distributions (Table 2) and the chosen range between -1.96 and 1.96 for both skewness and kurtosis.

|  | China |  |  |  |  | USA |  |  |  |  |
| --- | --- | --- | --- | --- | --- | --- | --- | --- | --- | --- |
|  | df (N-2) | rho | p-value | tau | p-value | df (N-2) | rho | p-value | tau | p-value |
| CO | 306 | .12 | **0.039** | .12 | 0.002 | 158 | .23 | .004 | .13 | **.015** |
| NO<sub>2</sub> | 306 | .73 | **0.000** | .54 | 0.000 | 247 | .63 | .000 | .45 | **.000** |
| O<sub>3</sub> | 306 | -.05 | 0.410 | -.03 | 0.442 | 752 | -.06 | .076 | -.11 | .000 |
| SO<sub>2</sub> | 306 | .61 | 0.000 | .40 | **0.000** | 315 | .02 | .661 | -.06 | .139 |

### 3.2 Correlation between air pollution variables with COVID-19 infections and mortality

Significant positive correlations between COVID-19 **infections** and air quality variables were found in each country (Table 4 and Graph 1 for NO_2_). In China, the strongest correlation was given by the satellite-derived CO values while in Italy and the U.S.A., the highest values were those of NO_2_ from satellite and ground measures, respectively. In general, Carbon Monoxide (CO), Formaldehyde (HCHO) and Nitrogen Dioxide (NO_2_) were positively correlated with COVID-19 cases, as well as particulate matter, especially PM2.5. Aerosol Index and SO_2_ show ambiguous behaviours, sometimes negatively correlated, and other times not significantly correlated. Likewise, Ozone shows a relatively strong positive correlation in Italy while it is negative in China and the U.S.A.

**Table 4.** Correlation between COVID-19 cases per 100,000 inhabitants and air quality variables.

| <u>cases_100k</u> | China |  |  | Italy |  |  | USA |  |  |
| --- | --- | --- | --- | --- | --- | --- | --- | --- | --- |
|  | df (N-2) | tau | p-value | df (N-2) | tau | p-value | df (N-2) | tau | p-value |
| Aerosol_sat | 345 | .01 | .797 | 105 | -0.1 | .000 | 3106 | -.12 | .000 |
| CO_sat | 345 | .34 | **.000** | 105 | 0.15 | **.024** | 3106 | .14 | **.000** |
| HCHO_sat | 345 | .13 | **.001** | 105 | 0.18 | **.007** | 3106 | .04 | **.006** |
| NO<sub>2</sub>_sat | 345 | .12 | **.004** | 105 | 0.52 | **.000** | 3106 | .20 | **.000** |
| O<sub>3</sub>_sat | 345 | -.16 | .000 | 105 | 0.35 | **.000** | 3106 | -.06 | .000 |
| SO<sub>2</sub>_sat | 345 | -.14 | .001 | 105 | -0.1 | .323 | 3106 | .00 | .863 |
| PM2.5_ground | 302 | .13 | **.003** | 88 | 0.31 | **.000** | 428 | .08 | **.014** |
| PM10_ground | 302 | .01 | .833 | 99 | 0.13 | .062 | 201 | .14 | **.004** |
| CO_ground | 302 | -.04 | .380 | - | - | - | 158 | .16 | **.004** |
| NO<sub>2</sub>_ground | 302 | .07 | .091 | - | - | - | 247 | .33 | **.000** |
| O<sub>3</sub>_ground | 302 | -.02 | .683 | - | - | - | 752 | -.06 | .009 |
| SO<sub>2</sub>_ground | 302 | -.06 | .173 | - | - | - | 315 | -.17 | .000 |

The **mortality** rate shows similar results to the COVID-19 infections (Table 5).

**Table 5.** Correlation between mortality rate and air quality variables (the mortality data for Italy were not publicly available at the comparable level of detail at the time of writing).

| Mortality | China |  |  | USA |  |  |
| --- | --- | --- | --- | --- | --- | --- |
|  | df (N-2) | tau | p-value | df (N-2) | tau | p-value |
| Aerosol_sat | 345 | .05 | .276 | 3106 | -.07 | .000 |
| CO_sat | 345 | .18 | .000 | 3106 | .09 | .000 |
| HCHO_sat | 345 | .16 | .000 | 3106 | .03 | .030 |
| NO<sub>2</sub>_sat | 345 | .10 | .019 | 3106 | .19 | .000 |
| O<sub>3</sub>_sat | 345 | .01 | .885 | 3106 | -.03 | .041 |
| SO<sub>2</sub>_sat | 345 | .00 | .933 | 3106 | .01 | .532 |
| PM2.5_ground | 302 | .19 | .000 | 428 | .14 | .000 |
| PM10_ground | 302 | .14 | .002 | 201 | .13 | .017 |
| CO_ground | 302 | .12 | .006 | 158 | .26 | .000 |
| NO<sub>2</sub>_ground | 302 | .12 | .005 | 247 | .27 | .000 |
| O<sub>3</sub>_ground | 302 | -.04 | .396 | 752 | -.06 | .022 |
| SO<sub>2</sub>_ground | 302 | .07 | .097 | 315 | -.10 | .022 |

In fact, there is a clear positive correlation with air quality variables, in particular with PM2.5 and CO in China, and with CO and NO_2_ in the U.S.A.

### 3.3 COVID-19 cases and air quality maps

The COVID-19 and air pollution maps for China were drawn with 10 and 6 classes, respectively. In the first case, we had to manually set the thresholds for the classes in order to obtain the best result. Due to the very large population and an apparently effective policy for the containment of the virus, the number of cases per 100,000 residents were relatively low and highly concentrated in the epicentre of the outbreak (the prefectures in the Hubei province).

The CO map was drawn using the equal count classification method (same number of features in each class). A visual correlation between the two maps can be perceived, especially between the eastern and western parts of the country.

**Figure 2.**
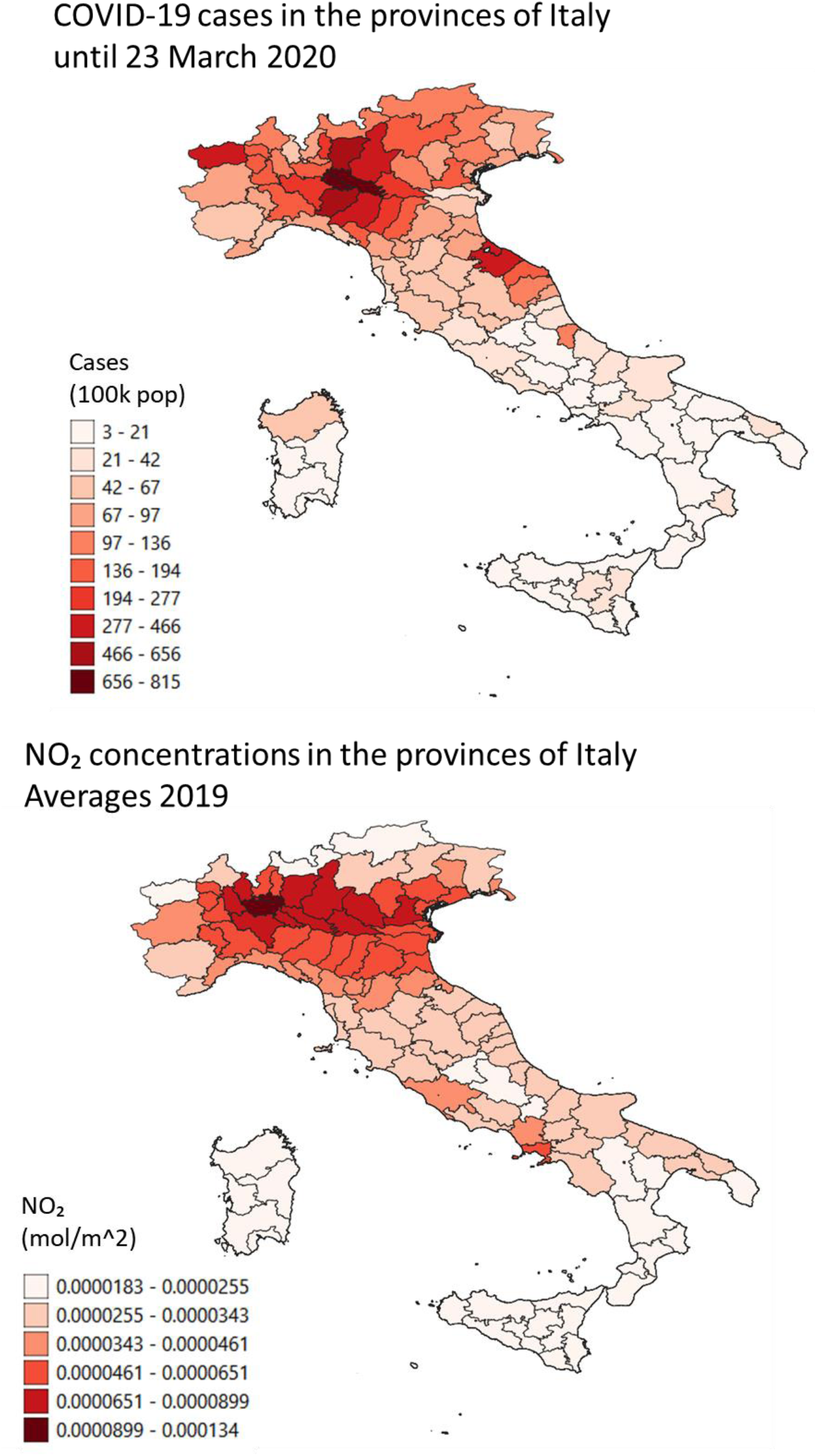
Map of COVID-19 total cases (controlled for population density) and NO_2_ concentrations in Italy.

Both maps for Italy were drawn by using a Natural Breaks classification method still with 10 and 6 classes, respectively, which highlights the heaviest affected areas by COVID-19 and NO_2_in the northern regions.

**Figure 3.**
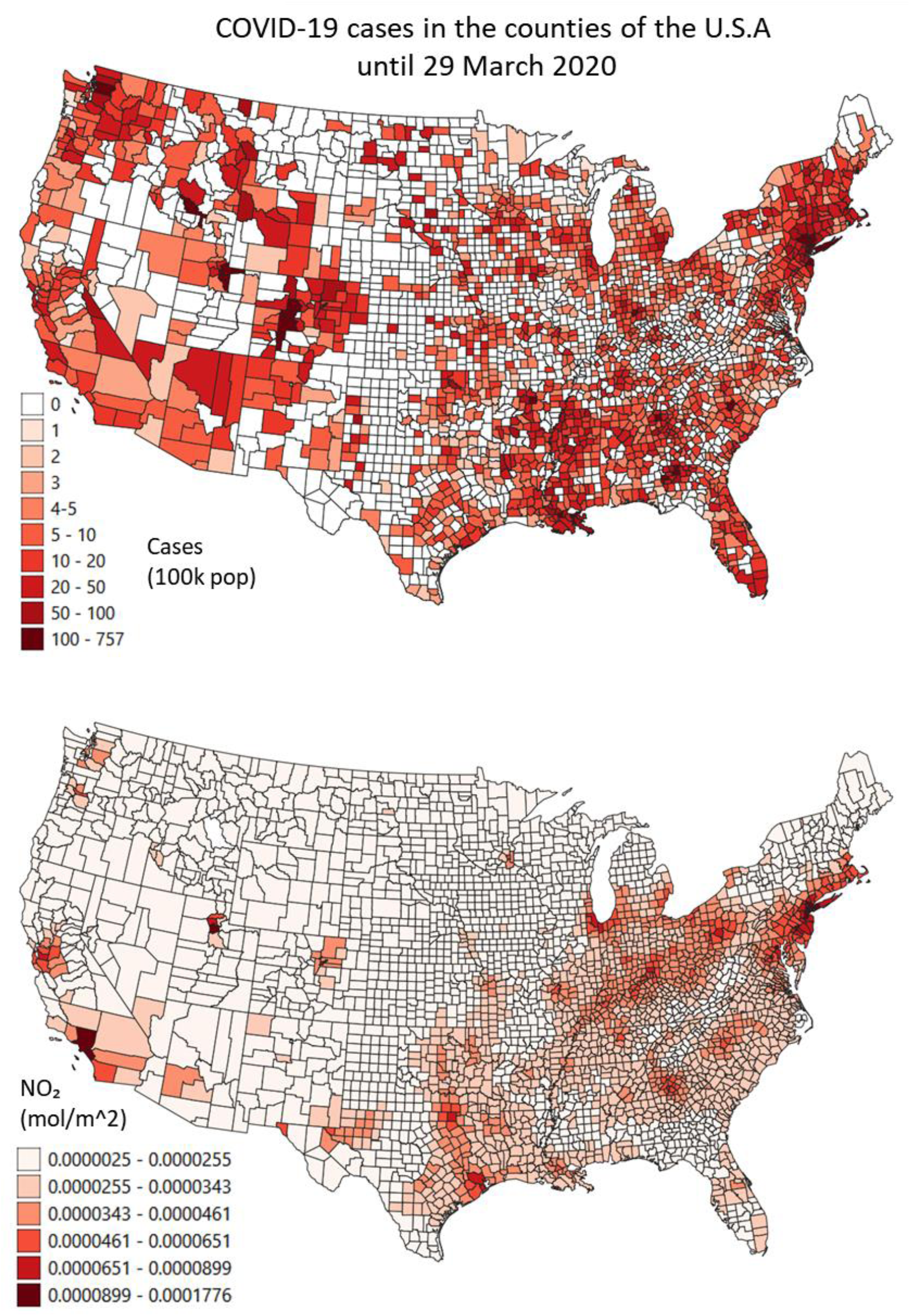
Map of COVID-19 total cases (controlled for population density) and NO2 concentrations in the U.S.A.

The U.S.A. COVID-19 maps respect the classification method employed for China. Here the virus noticeably appears more widespread over the country’s territory. The highest values are found in more polluted areas. The NO_2_ map, instead, follows the classification method used in Italy.

By comparing the three countries, it seems that Italy faces a more critical problem of NO_2_ pollution during the year 2019, relative to its size.

## 4. Discussion

This study is the first to empirically investigate air pollution for three countries as a potential risk factor for the incidence and mortality rates of COVID-19. It provides preliminary evidence that COVID-19 cases are most often found in highly polluted areas of China, Italy and the United States regardless of population density. In addition, in these areas of low air quality, the virus kills more often than elsewhere.

The interpretation of these findings has to be necessary **cautious**, as the virus spread especially in Italy and the United States is still ongoing^68^ and is being contained^48^. However, it is remarkable that we obtained consistent significant correlations between air pollution variables and risk factors for COVID-19, despite varying population densities within and between the countries. By controlling for the number of infections per 100,000 habitants, we found statistically significant, positive correlations between COVID-19 infections and low air quality in each of the three assessed countries.

In 2020, in China, in Italy and in the Unites States we found more **infections** in those areas afflicted by Carbon Monoxide, formaldehyde, Nitrogen Dioxide, and PM2.5. In Italy, also the Ozone variable was strongly correlated. In Hubei province, a brand new time analysis gives preliminary evidence of a correlation between NO_2_ high levels and 12-day delayed virus outbreaks.

Infected people were more likely to die in the Chinese, Italian, and American areas with poor air quality, regardless of the higher number of cases. Our **mortality** ratio was higher and in particular so in China, where PM2.5 and CO were at levels considered unhealthy, and in the U.S.A. where CO and NO_2_ were higher than the average. In the U.S.A., levels of PM2.5 have just been found responsible for 20-time higher mortality rate by COVID-19, a rate much higher than other demographic co-variables^69^. We found that in Italy, the correspondence between poor air quality and SARS-CoV-2 appearance and its induced mortality was the starkest. It should be noted that the Italian higher mortality than the one predicted from mathematical modelling is unlikely caused by genetic mutations of the virus^70^. Therefore, other factors must be attributed to such a stronger virulence. We still need to obtain the Italian mortality data at the small administrative level, but pollution seems to be one of the suitable predictors.

We were able to validate this methodology, since the **series of variables** we took into account were correlated with each other. In particular, Carbon Monoxide (CO), Formaldehyde (HCHO) and Nitrogen Dioxide (NO_2_) were correlated, as well as particulate matter, especially PM2.5. Satellite NO_2_ and CO are suitable representatives for general air quality^71^, since they are more consistent than ground station data, providing finer detail, consistency, accuracy, with virtually no errors of counterfeits. They are also to a certain extent less prone to biases from weather conditions^58^ of wind and greenhouse effect of temperature inversion, in turn also related to air pollution.

We used annual means, which only partially represent the real emission of pollutants during the year and do not make evident seasonal variations and other fluctuations. However, our aim was to highlight differences in air quality within a country’s region and show the correlation with the virus. Therefore, threshold values of air pollution cannot be inferred from this study.

Since we have now some first evidence that the cross of the virus from animals to humans may have happened years earlier than the end of 2019 in the Chinese city of Wuhan^44^, we can speculate that air pollution could have played a role in gradually **exacerbating morbidity and mortality**, mutating the virus from an initial evolutionary stage not causing any more serious morbidity than a cold, to becoming so threatening to humans.

In the unlikely case that the figures provided by the states in relation to number of infections and deaths are inaccurate^72^, our analyses and conclusions would not need to be reframed. If that is the case, this error will most likely be concentrated in just one or very few administrations and it would not affect our very large correlational dataset.

The possibly large proportion of **asymptomatic cases** has been implied as an important factor in the fast spread of the virus and will necessarily lead to a biased mortality rate. Different government policies with regards to testing have led to vastly different estimates across countries^73,41^, and a COVID-19 overall mortality rate has not been established yet. Asymptomatic cases could be as high as about 50% of total cases, as estimated by simulations^74^ or recorded in Iceland^75^, where mass screening with oropharyngeal tests^76^ was employed.

States have responded by trying to mitigate the spread of the virus through imposing widespread lockdowns. This has led to a **decrease in air pollution**, which in China likely prevented the deaths of 4,000 children under 5 and 73,000 adults over 70^77^. However, the winter months and low temperature caused people to keep the heating systems on, maintaining a certain amount of pollution (coal and electricity in China, gas methane in Italy, gas and electricity in the United States). In Europe^78^ and in China^79^ a consistent reduction in air pollution was recorded by satellites due to reduced anthropogenic activities during the lockdowns, although it occurred gradually^80,81^, also due to weather conditions unfavourable to air quality. The quarantines certainly decreased the role that commuting has in the virus spread. Nonetheless, reduced anthropogenic activities and reduced mobility lose correlational significance over time, after the first stages of the infection^55^. Instead, the correlation with low air quality remains significant throughout the different epidemic stages.

This pandemic has not ended yet, so our conclusions are necessarily **restricted** to the stage of infection of those three countries. There are also confounding factors such as how the virus infection was determined in patients by different countries. However, the larger the geographical areas that are affected by the pandemic, the lower these elements should play a role.

Our study is eventually going to be completed with the analysis of an up to date dataset and possibly additional countries, before its formal journal submission. **Further research** in the field of physics should also be endorsed to investigate the capacity of air pollutants to act as viral vectors. Air pollutants may in fact act as a medium for the aerial transport of SARS-CoV-2^32^, potentially broadening the harm done in the contagions.

Our results inform epidemiologists on how to prevent future, possibly more lethal viral outbreaks by curbing air pollution and climate change. Institutions need to endorse such **interventions** more seriously together with other comprehensive measures playing a role in reducing epidemics, such as impeding biodiversity loss, ending wars, and alleviating poverty^82^.

## Data Availability

The data are going to be available online once peer-reviewed by a journal.

https://github.com/DavideFornacca

## Acknowledgments

Lei Shi and Xiao Wen commented the study. Livia Ottisova improved and revised the manuscript.

## Data Availability

Data available online once peer-reviewed by a journal.

